# Use of the International Classification of Functioning, Disability and Health (ICF) in Randomized Controlled Trials of Rheumatoid Arthritis Pharmacological Treatments: a Scoping Review

**DOI:** 10.1101/2025.07.14.25331488

**Authors:** Adrian Martinez-De la Torre, Polina Leshetkina, Ogie Ahanor, Roxanne Maritz

## Abstract

**Background:** Rheumatoid arthritis (RA) is a chronic autoimmune disease affecting approximately 0.5–1.0% of the adult population and is a significant contributor of disability worldwide. While Phase III randomized controlled trials (RCTs) remain the gold standard for evaluating pharmacological treatments, they often fail to capture outcomes that reflect patients’ lived experiences, also referred to as functioning. The International Classification of Functioning, Disability and Health (ICF) Brief Core Set for RA offers a standardized, patient-centered framework for assessing functioning across relevant domains.

**Objective:** To examine the extent to which functioning-related outcomes in Phase III pharmacological RCTs for RA align with the ICF Brief Core Set for RA and to identify the most frequently represented functioning categories.

**Methods:** A scoping review was conducted following PRISMA-ScR guidelines. Literature was searched in MEDLINE, EMBASE, and ClinicalTrials.gov from 2010 to 2025. Phase III RCTs evaluating pharmacological interventions in adult patients with RA were included. Functioning-related outcomes were extracted and mapped to ICF categories using standardized linking rules.

**Results:** Of 852 records screened, 91 met the inclusion criteria. Functioning was frequently assessed through patient-reported outcomes and composite clinical measures. The most commonly linked ICF categories included sensation of pain (b280) and mobility of joint functions (b710) from the body functions domain; walking (d450) and carrying out daily routine (d230) from the activities and participation domain; and structures of the shoulder (s720), upper (s730), and lower extremities (s740) from the body structures domain. However, none of the studies explicitly used the ICF framework.

**Conclusion:** Functioning is implicitly assessed in RA pharmacological trials, yet the ICF framework remains underutilized. Explicit integration of the ICF Brief Core Set for RA into trial design could improve the standardization, comparability, and patient-centeredness of outcome measurement, ensuring that clinical research better reflects what matters most to individuals living with RA.

## Introduction

Rheumatoid Arthritis (RA) is a chronic, progressive autoimmune disease that significantly contributes to global disability and associated with early mortality. It is estimated to affect between 0.5 and 1.0% of the adult population worldwide, increases in prevalence with age, and affects more women than men [1]. In 2020, approximately 17.6 million people were affected by RA worldwide, with projections estimating a rise to 31.7 million by 2050 [2]. The condition imposes a substantial economic and societal burden by limiting individuals’ functioning, contributing to loss of productivity, and increasing healthcare costs [3].

As the global burden of RA continues to grow, rigorous evaluation of pharmacological treatments has become increasingly important. Phase III randomized controlled trials (RCTs) remain the gold standard for assessing the efficacy and safety of new therapies. However, these trials are often conducted in highly controlled settings with narrowly defined patient populations and rely on predefined clinical endpoints. As a result, they often overlook treatment effects that directly impact patients’ daily lives, particularly regarding functioning and participation [4].

Functioning is increasingly recognized as the third health indicator, alongside mortality and morbidity, and is central to rehabilitation practice and policy [5]. To support this, in 2001 the World Health Organization (WHO) developed the International Classification of Functioning, Disability and Health (ICF) to provide a standardized framework for assessing functioning across health conditions [6,7]. The ICF integrates biological aspects, such as body functions and structures, and lived health, including activities and participation in context with environmental factors [8]. To tailor this framework to specific conditions, condition-specific Core Sets were developed [9]. The ICF Core Set for RA was developed in 2004, identifying the most relevant ICF categories for describing functioning in RA patients [10]. These Core Sets have been validated from both patient and clinician perspectives [11,12]. Moreover, the WHO Rehabilitation 2030 initiative further emphasizes the importance of integrating functioning into health systems globally, in line with Sustainable Development Goal 3 to promote health and well-being globally [13].

A growing body of literature in rheumatology has explored how clinical outcomes can be linked to the ICF framework. One systematic review mapped outcome measures from trials on joint protection programs and assistive devices in hand osteoarthritis and RA to relevant ICF categories [14]. Another examined both randomized trials and real-world studies of acute rheumatologic conditions, linking commonly used patient-reported outcomes (PROs) to the ICF Comprehensive Core Set for RA [15]. Additionally, another systematic review categorized factors associated with hand function in people with RA using the ICF framework [16].

While the ICF framework has been applied in studies of non-pharmacological interventions, it is still rarely applied in pharmacological clinical trials. Its close alignment with patient-reported outcome measures makes it a valuable tool for capturing aspects of treatment that matter most to patients. Integrating the ICF Core Set into pharmacological research could enhance understanding of drug safety, efficacy, and patient well-being, while also supporting more personalized and function-oriented care.

This review examines how functioning is assessed in RA clinical trials, evaluates the extent to which outcome measures can be linked to the ICF Brief Core Set for RA, and identifies the most frequently represented categories. The findings aim to inform the integration of the ICF into pharmacoepidemiological research and promote patient-centered approaches to treatment evaluation.

## Methods

The study design for this scoping review adheres to the Preferred Reporting Items for Scoping Reviews (PRISMA-ScR) guidelines [17] (Supplementary Table S1). This approach was chosen to systematically map existing evidence of the ICF Brief Core Set for RA categories usage in Phase III RCTs, identify gaps in its application, and explore opportunities for the integration of functioning outcomes into clinical trials. The protocol for this review was registered with the Open Science Framework (OSF) on 24 February 2025, with the registration reference available at https://doi.org/10.17605/OSF.IO/SX9AW.

### Eligibility criteria

The inclusion and exclusion criteria for eligible studies are outlined in Table 1. We focused on phase III RCTs evaluating pharamacological interventions for RA, as these trials represent the critical stage in the clinical development process where therapeutic efficacy and safety are assessed in larger, more heterogeneous patient populations. Additionally, we used the ICF Brief Core Set for RA because it was specifically developed to guide outcome assessment in both clinical trials and routine clinical practice. It captures the most essential functioning categories relevant to individuals with RA, based on a comprehensive, consensus-driven process involving patients, clinicians, and experts [18]. The time frame for this scoping review was selected starting from 2010, coinciding with the publication of the updated classification criteria by the American College of Rheumatology (ACR) and the European League Against Rheumatism (EULAR), which introduced more accurate diagnostic approaches and more effective treatment strategies for RA [19].

**Table 1.** Eligibility criteria.

| INCLUSION | EXCLUSION |
| --- | --- |
| Phase III RCTs | Phase I and II RCTs |
| Study must investigate pharmacological intervention for RA | Study investigates non-pharmacological intervention for RA (i.e., alternative therapies, assistive technologies, diet, exercise, rehabilitation and surgical treatment) |
| Functioning outcomes are assessed (matching ICF categories according to ICF Brief Core Set for RA) | RCTs that assess only drugs safety, immunogenicity, pharmacodynamic and pharmacokinetic outcomes |
| Adult participants with the diagnosis of RA, either according to ACR criteria or 2010 ACR/EULAR criteria | Pediatric participants with the diagnosis of RA |
| Full study is available | RWE (observational i.e., cohort, cross-sectional studies) |
| Study must be published from 2010 to February 2025 | Languages other than English (for full text) |

### Information sources and search strategy

A literature search was performed in MEDLINE via Ovid®, EMBASE®, and ClinicalTrials.gov® from 2010 to February 2025. Medical subject headings (MeSH) and free text keywords were used to develop the search strategy, which was formulated in MEDLINE via Ovid and adapted for usage in other databases. Published search filters were applied to restrict the results to Phase III pharmacological RCTs in adult populations. Full search strategies are detailed in Table 2.

**Table 2.**
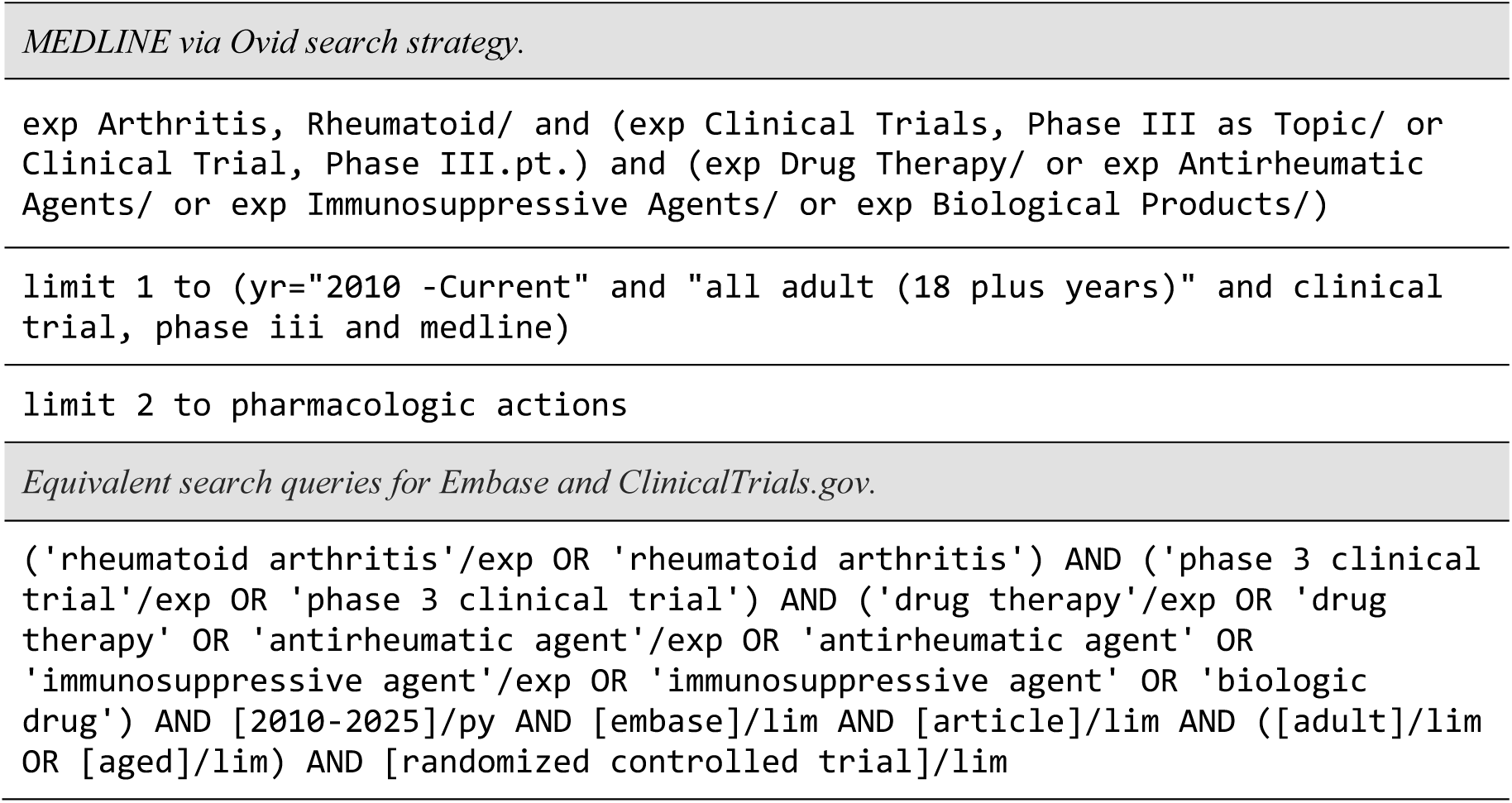
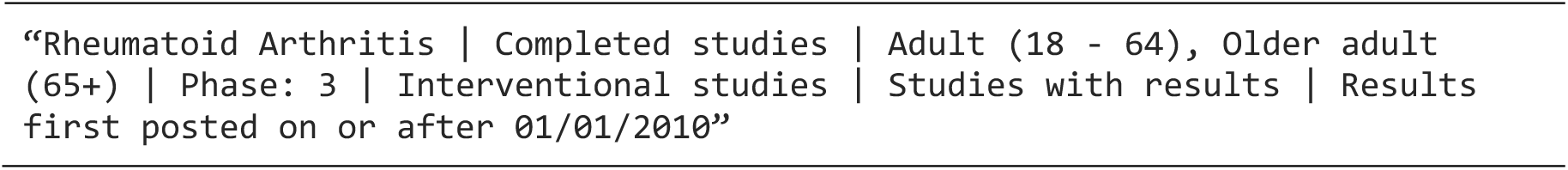
Search strategies used to identify Phase III pharmacological trials in rheumatoid arthritis across MEDLINE, Embase, and ClinicalTrials.gov (2010–2025).

### Study selection and data collection

All studies identified through database searches that met inclusion criteria were imported into Zotero® (6.0.37/2024) library for data management. After duplicate removal using Covidence®, the study selection process was conducted in two stages.

In the first stage, titles and abstracts were screened according to the eligibility criteria outlined in Table 1. When titles and abstracts lacked sufficient information to determine eligibility, the articles were retained for full text screening.

In the second stage, the full texts of the identified publications were retrieved and reassessed against the eligibility criteria to determine study inclusion for data extraction.

### Data extraction and critical appraisal

Information extraction occurred in two steps by a primary reviewer (PL). First, publication characteristics, including author and year, title, drug names, countries, and total number of participants, were uniformly extracted from each paper by Covidence.

In the second step, all functioning-related outcomes were extracted alongside clinical efficacy, safety, immunogenicity, pharmacodynamics, and pharmacokinetics outcomes. Secondary endpoints, including clinical efficacy measures, were further categorized into PROs, clinical and technical measures to facilitate the analysis. Environmental factors (5 categories) were not assessed in this review due to their limited relevance and representation in the design and reporting of Phase III pharmacological RCTs.A second reviewer (AMDLT) provided oversight and verified the extracted data for consistency and completeness.

### Data analysis and presentation

A PRISMA-ScR flowchart illustrates the flow of information through the scoping review’s phases. The absolute and relative frequencies of various outcome assessment tools used in RCTs were calculated and summarized in the table.

The concepts in outcome assessment tools were linked to the ICF categories represented in theBrief Core Set for RA using standardized linking rules (2019) [20]. When a concept was too broad to be fully captured by a single ICF category, it was linked to the most appropriate broader category, captured by the Core Set. For example, the FACIT-F item “I have trouble walking long distances because of my fatigue” was linked to walking (d450). The ICF Brief Core Set categories that could not be linked to any concept described by an assessment tool were also documented as a gap in the table. A heatmap was created to visualize how the ICF categories were addressed by the outcome assessment tools in the included studies.

## Results

### Study selection

The search of the selected databases resulted in the retrieval of 852 citations (MEDLINE via Ovid: n=285; EMBASE: n=326; ClinicalTrials.gov: n=241). After removing duplicates, 685 articles remained for screening. Of these, 594 were excluded based on the eligibility criteria. The selection process is detailed in the PRISMA-ScR flowchart (Figure 1). Ultimately, 91 articles met all inclusion criteria and were included in the final review.

**Figure 1.**
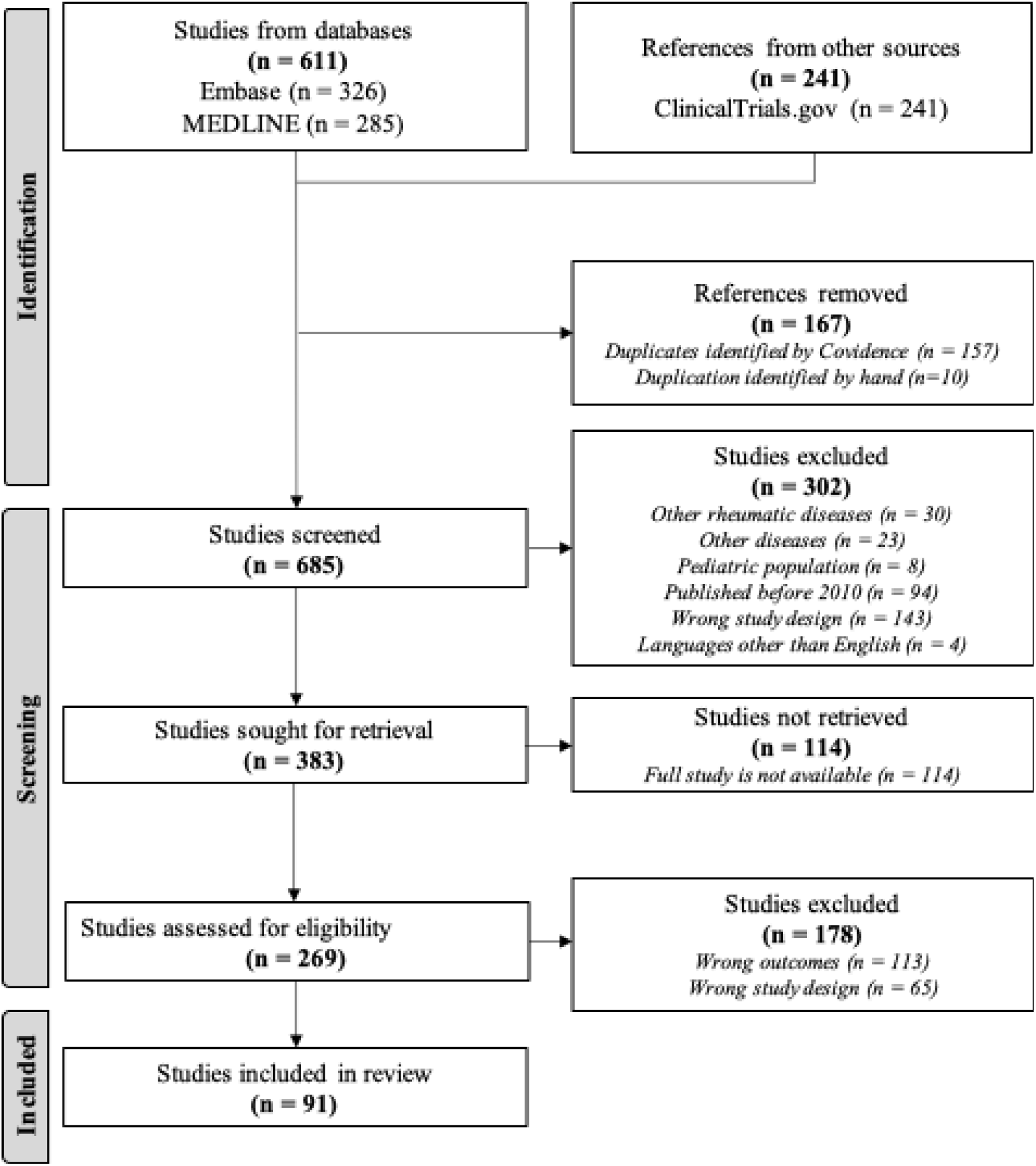
PRISMA-ScR flow diagram illustrating the study selection process.

### Characteristics of included studies

The 91 studies included in this review were Phase III RCTs evaluating pharmacological interventions for any form of RA. These trials varied in duration and were predominantly multicenter, often conducted across multiple countries, with the majority taking place in Europe and the United States. Sample sizes varied, with some studies including over 1000 participants, while others had around 150 to 500 participants. The studies included both monotherapy and combination therapy approaches, mostly targeting patients with moderate-to-severe active rheumatoid arthritis who had inadequate responses to previous treatment strategies. RCTs covered a broad range of drug classes, predominantly Disease-Modifying Antirheumatic Drugs (DMARDs), including biologic DMARDs (bDMARDs), traditional DMARDs (tDMARDs), and synthetic DMARDs (sDMARDs). Study characteristics are summarized in Supplementary Table S2.

### Assessment of functioning-related outcomes in RA Phase III trials

Among the primary endpoints, clinical efficacy outcomes, such as the American College of Rheumatology response (ACR20) and Disease Activity Score (DAS-28), were reported in 54.9% and 52.8% of studies, respectively, as shown in Table 3. All RCTs evaluated safety outcomes, such as Adverse Events (AEs), as primary or secondary outcomes.

**Table 3.** Absolute and relative frequencies of outcome assessment tools used across included RCTs.

| Measures | Number of studies | Percentage (%) |
| --- | --- | --- |
| **Primary Clinical Efficacy Outcomes** |  |  |
| ACR20 response criteria | 50 | 54.9 |
| DAS-28 response criteria/EULAR response criteria | 48 | 52.8 |
| **Secondary Efficacy Outcomes (multiple measures per each RCT)** |  |  |
| **Clinical Measures** |  |  |
| DAS-28/DAS-28(CRP)/DAS-28(ESR) | 78 | 85.7 |
| ACR20/50/70 | 66 | 72.5 |
| CDAI/SDAI | 40 | 43.9 |
| PhGA (measured by VAS mm) | 18 | 19.8 |
| MS | 10 | 10.9 |
| TJC/SJC | 10 | 10.9 |
| **Patient-Reported Outcomes** |  |  |
| Physical function (measured by HAQ) | 61 | 67.0 |
| Health-related quality of life (measured by SF-36 PCS and MCS) | 43 | 47.3 |
| Pain (measured by VAS) | 22 | 24.2 |
| Fatigue (measured by FACIT-F) | 22 | 24.2 |
| PGA (measured by VAS mm) | 12 | 13.2 |
| Health-related quality of life (measured by EQ-5D-5L) | 8 | 8.8 |
| Working limitations (measured by WPS/WPAI-RA) | 6 | 6.6 |
| Sleep (measured by MOS-Sleep scale) | 4 | 4.4 |
| **Technical Measures** |  |  |
| CRP | 55 | 60.4 |
| ESR | 36 | 39.6 |
| ADA | 34 | 37.4 |
| X-ray (Sharp score/Modified, Sharp/van der Heijde score/different joints, score not specified) | 19 | 20.9 |

Note: ACR American College of Rheumatology, DAS-28 Disease Activity Score with the 28-joint count, EULAR European League Against Rheumatism, SDAI Simplified Disease Activity Index, CDAI Clinical Disease Activity Index, PhGA Physician Global Assessment of Disease Activity, MS Morning Stiffness, TJC and SJC Tender Joints Count and Swollen Joint Count, HAQ Stanford Health Assessment Questionnaire, SF-36 Short Form 36 with physical health summary (PCS) and the mental health summary (MCS), VAS Visual Analogue Scale, FACIT-F Functional Assessment of Chronic Illness Therapy – Fatigue Scale, PGA Patient global assessment of disease activity, WPS Work Productivity Survey, WPAI-RA Work Productivity, and Activity Impairment for Rheumatoid Arthritis, MOS-sleep scale Medical Outcomes Study Sleep Scale, CRP C-reactive protein, ESR erythrocyte sedimentation rate, ADA anti-drug antibody.

Secondary endpoints were grouped into clinical, patient-reported, and technical measures. C*linical measures* refer to physician-assessed indicators used to evaluate disease activity, severity, and treatment responses. ACR20/50/70 response thresholds were reported in 72.5% of studies, extending beyond their role as primary endpoints. DAS-28 and its variants incorporating C-Reactive Protein (CRP) or Erythrocyte Sedimentation Rate (ESR) were more frequently used, appearing in 85.7% of studies. Simplified and Clinical Disease Activity Index (SDAI/CDAI) were reported in 43.9% of RCTs. In 10.9% of studies, Tender and Swollen Joint Counts (TJC/SJC) were assessed as separate measures, beyond their inclusion in composite clinical indices.

*PROs* refer to patient reported outcome measures that reflect the patients’ perspective on their health status, functioning, and well-being. In 13 RCTs, PROs were assessed as the primary endpoint. Physical function was assessed using the Health Assessment Questionnaire (HAQ) in 67% of RCTs separately from the ACR core set, which includes this assessment tool. Health-related quality of life was measured through the Short Form-36 in 47.3% and through EQ-5D-5L in 8.8% of studies. Less frequently included tools were pain measurement using a Visual Analog Scale (VAS) in 24.2%, measures of fatigue through Functional Assessment of Chronic Illness Therapy – Fatigue Scale (FACIT-F) in 24.2%, Patient Global Assessment of disease activity (PGA) in 13.2%, work limitations with Work Productivity Survey (WPS) and Work Productivity and Activity Impairment (WPAI) in 6.6% of studies.

*Technical measures* refer to objective assessments based on laboratory tests or imaging used to monitor biological activity and disease progression. Inflammatory markers were frequently assessed separately from DAS-28, with CRP reported in 60.4% and ESR in 39.6% of the studies. Radiographic assessments, including all X-Ray scoring methods, were applied in 20.9% of trials that ran longer than 12 months.

### Linking outcomes assessed in RA RCTs to the ICF Brief Core Set for RA

The ICF Core Set for RA includes 23 categories across 4 domains: body functions (5 categories), activities and participation (6 categories), and body structures (4 categories). Table 4 presents the linkage between the ICF Brief Core Set for RA categories and the functioning-related outcome measures identified across studies.

**Table 4.**
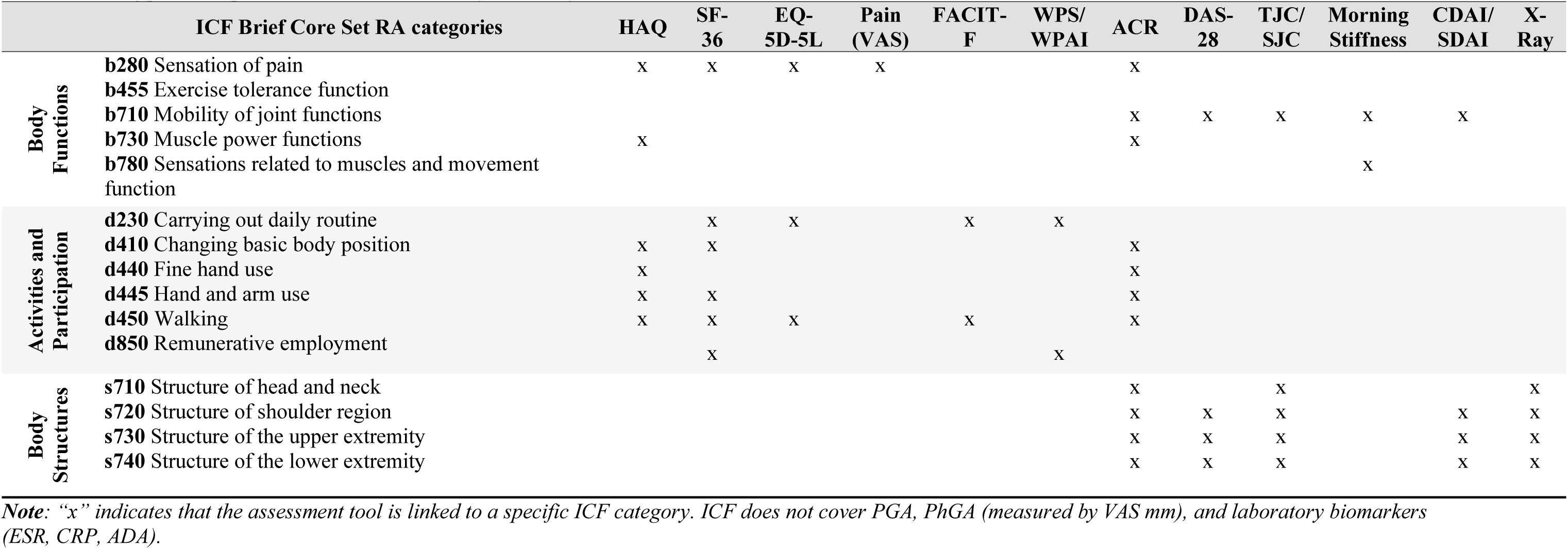
Linking patient-reported measures to ICF Brief Core Set for RA.

*PROs* predominantly covered categories from *activities and participation (d)* and *body functions (b)* domains of the ICF Brief Core Set. The HAQ and SF-36, previously linked to the ICF Comprehensive Core Set for RA by Stucki [10], also align with corresponding categories in the Brief Core Set. The EQ-5D-5L, which assesses mobility, self-care, usual activities, pain/discomfort, and anxiety/depression was linked to *sensation of pain (b280), carrying out daily routine (d230), and walking (d450)*. The pain visual analogue scale (VAS) was also linked to *sensation of pain (b280)*. FACIT-F, comprising 13 items capturing the impact of fatigue on daily activities, walking, and mood, was mapped to *carrying out daily routine (d230)* and *walking (d450)* categories. WPS and WPAI were primarily linked to *remunerative employment (d850)*, with WPS also addressing *carrying out daily routine (d230)* category.

*Sleep functions (b134)*, assessed through the Medical Outcomes Study - Sleep Scale (MOS-Sleep Scale) in 4 RCTs, and *emotional functions (b152)*, assessed through the SF-36 and FACIT-F, are categories from the ICF Comprehensive Core Set but are not included in the RA Brief Core Set.

*Clinical measures* were mainly linked to the ICF domains of *body functions (b) and body structures (s).* TJC/SJC were mapped to *mobility of joint functions (b710)* and all categories within *body structures* domain. The ACR response criteria, which include TJC-68, SJC-66, HAQ, PGA, PhGA, pain scale, and CRP/ESR, were linked to *mobility of joint functions (b710)* category, *body structures* domain, with the HAQ component linking to the *activities and participation* domain. Similarly, DAS-28 and CDAI/SDAI scores, which include TJC-28, SJC-28, PGA, and CRP/ESR were linked to *mobility of joint functions (b710)* and *structure of the shoulder region (s720), upper extremities (s730),* and *lower extremities (s740).* Morning stiffness (MS) was linked to the *mobility of joint functions (b710)* and *sensations related to muscles and movement function (b780)*.

X-Ray-based joint assessments, considered *technical measures,* were linked to all relevant *body structures* categories.

### ICF category representation in RA RCTs assessment tools

Figure 2 presents a heatmap illustrating how frequently outcome assessment tools used in Phase III RA trials addressed specific categories from the ICF Brief Core Set for RA. The horizontal axis lists the tools, while the vertical axis shows the 15 ICF categories to which they were linked. This visualization highlights the extent to which each domain of functioning is represented in pharmacological trials and reveals important gaps.

**Figure 2.**
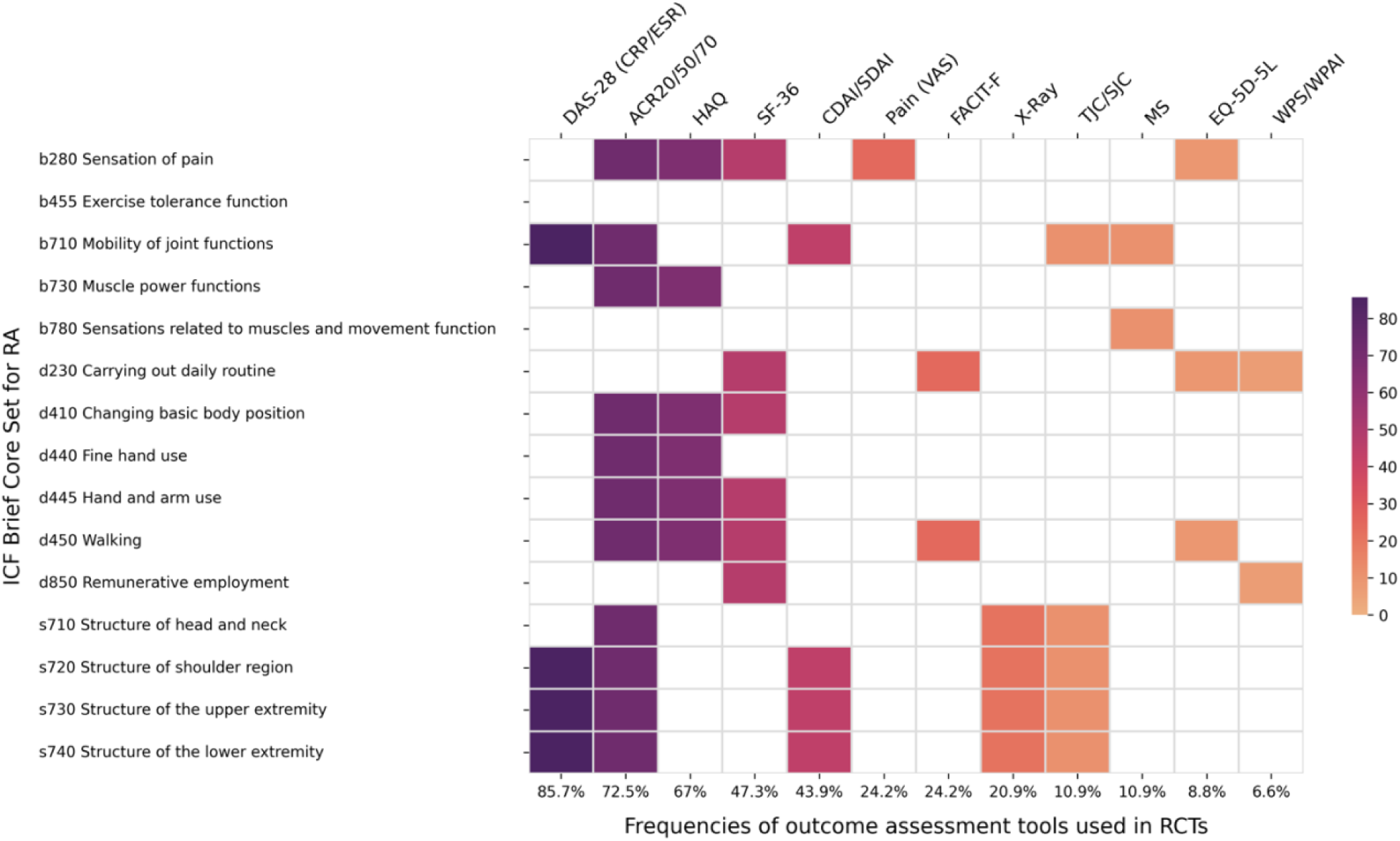
Heatmap of relevant frequencies of outcome assessment tools used across RA RCTs and corresponding ICF Brief Core Set categories. Note: Each cell in the heatmap indicates how frequently a specific outcome assessment tool (shown on the horizontal axis) was linked to a particular ICF category (listed on the vertical axis) across the included RCTs. Color intensity reflects frequency, with darker shades representing higher usage. The percentage values beneath each column indicate the proportion of studies in which that outcome tool was used.

Within the body functions domain, *sensation of pain* (b280) and *mobility of joint functions* (b710) were the most frequently addressed categories. These were consistently captured by a range of tools, including ACR, HAQ, SF-36, EQ-5D-5L, and pain scales. In contrast, *exercise tolerance functions* (b455) were not assessed by any tool, and *muscle power functions* (b730) appeared only in the HAQ and ACR, indicating limited attention to fatigue-related impairments in trial outcomes.

In the activities and participation domain, *walking* (d450) and *carrying out daily routine* (d230) were the most frequently represented, particularly through PROs such as the HAQ, SF-36, and FACIT-F. Categories related to fine motor function, including *changing basic body position* (d410), *fine hand use* (d440), and *hand and arm use* (d445), were less commonly addressed and primarily captured by HAQ and SF-36. *Remunerative employment* (d850) was assessed in 47.3% of trials using SF-36 and in 6.6% using WPS/WPAI, reflecting limited but notable consideration of work-related functioning.

The body structures domain showed the highest overall coverage. Categories such as *structure of the shoulder region* (s720), *upper extremity* (s730), and *lower extremity* (s740) were evaluated in all trials through joint counts (TJC/SJC) and in 20.9% of studies via radiographic assessments. These findings underscore the strong emphasis placed on structural joint integrity in RA pharmacological trials, compared to more variable attention given to broader aspects of functioning.

## Discussion

This review examined how functioning is assessed in Phase III RCTs for rheumatoid arthritis and whether these assessments align with the ICF Brief Core Set for RA. By identifying and mapping functioning-related outcomes to ICF categories, we found that many commonly used assessment tools correspond closely to the domains of the Brief Core Set. However, none of the included trials explicitly used the ICF as a guiding framework for outcome measurement.

Functioning is increasingly recognized as a core indicator of health, complementing traditional metrics such as mortality and morbidity by capturing what truly matters to people in their daily lives [21]. In rheumatologic conditions, patients experience higher morbidity and mortality than the general population, largely due to systemic inflammation, cardiovascular complications, and comorbidities [22]. However, beyond these clinical endpoints, RA often leads to substantial and persistent limitations in functioning that directly affect patients’ well-being. Pain, fatigue, reduced mobility, and limitations in work or social participation can persist even when clinical or biological markers improve [23]. This underscores the importance of systematically integrating functioning into clinical trials using a standardized framework like the International Classification of Functioning, Disability and Health (ICF) [8]. Functioning outcomes should be considered as primary endpoints in RA trials, not secondary or exploratory, because improvements in biomarkers alone may fail to reflect meaningful changes in how patients feel, function, and participate in everyday life.

Our findings contribute to a growing recognition of the importance of incorporating functioning as a key outcome in clinical trials, particularly in contexts such as rheumatoid arthritis (RA), where traditional biomarkers or mortality outcomes may not fully capture the burden of disease. Functioning reflects what matters most to patients, how they feel and perform in their daily lives, and offers a crucial complement to morbidity and mortality [5]. The ICF provides a standardized and person-centered framework for assessing functioning across health conditions and contexts. While the ICF has been extensively adopted in rehabilitation medicine to guide clinical assessment and care planning, for example, in spinal cord injury and stroke, its application in pharmacological trials remains limited [24,25].

Although the ICF is not mentioned explicitly in RA Phase III trials, our findings showed that many commonly used outcome measures align closely with categories in the ICF Brief Core Set for RA, indicating that functioning is already being assessed implicitly. For example, *mobility of joint functions* (b710) and *sensation of pain* (b280) were addressed in over 85% of studies through tools such as ACR, DAS-28, HAQ, and pain scales. The HAQ, used in 67% of RCTs, captured several categories from the activities and participation domain, including *carrying out daily routine* (d230) and *walking* (d450). Similarly, SF-36, included in nearly half of the trials, was linked to categories spanning both body functions and participation, such as *emotional functions* (b152) and *remunerative employment* (d850). Despite this substantial overlap, the ICF framework itself was never used to guide outcome selection.

Making this linkage explicit would enhance the consistency, comparability, and relevance of outcome assessments across trials. Recognizing the ICF as a unifying framework would also support the development of standardized functioning outcomes, which can bridge clinical efficacy with real-world impacts on patients’ lives. Ultimately, centering functioning in RA trials reinforces a person-centered approach that values outcomes meaningful to patients and supports more holistic treatment evaluations.

Several methodological limitations should be acknowledged. First, data extraction and screening were primarily conducted by a single reviewer, which introduces a risk of selection and classification bias. Although a second reviewer supervised the process and verified the included references, they were not involved in the initial extraction, which may limit the reliability of decisions regarding study inclusion and ICF linking. Second, the review focused exclusively on Phase III RCTs of pharmacological interventions for rheumatoid arthritis, excluding earlier-phase trials and studies of non-pharmacological or adjunctive therapies. This narrow scope may have resulted in the omission of relevant data on functioning outcomes used in other trial types. Third, trials that focused solely on technical outcomes (e.g., imaging or laboratory-based biomarkers) or safety endpoints without including functioning-related measures were excluded, which may have introduced outcome reporting bias. Finally, the wide variability in how outcome assessment tools were named, formatted, or reported across studies could have led to inconsistencies in classification or underestimation of specific functioning domains. While we applied standardized ICF linking rules by Cieza and colleagues [20], the process still required interpretation and may have introduced a limited element of subjectivity.

Our work presents several notable strengths. It is the first study to systematically examine the extent to which functioning-related outcomes in Phase III RCTs for rheumatoid arthritis (RA) align with the ICF framework, specifically the ICF Brief Core Set for RA. By mapping commonly used assessment tools to ICF categories, the review offers a novel, structured understanding of how functioning is implicitly captured in RA trials, despite the ICF framework not being explicitly applied. The study also highlights important gaps in the representation of key functioning domains, such as activities and participation, providing a clear rationale for their more systematic inclusion in future trial designs. The methodological rigor of this review was further strengthened by adherence to PRISMA-ScR guidelines, the use of established ICF linking rules, and the comprehensive review of multiple data sources (MEDLINE, Embase, ClinicalTrials.gov), supporting the reproducibility of findings. By bridging clinical outcomes with functioning categories, the study contributes valuable insights that can inform more patient-centered and comprehensive approaches to outcome assessment in rheumatology.

The findings of this review have important implications for health research policy and regulatory frameworks. There is a clear need to systematically include functioning outcomes, grounded in the ICF framework, in the design and evaluation of pharmacological clinical trials. In the context of RA, where biomarkers may not always reflect the lived experience or long-term impact of the disease, integrating functioning outcomes ensures that trials capture dimensions of health that truly matter to patients. Recognizing the ICF Brief Core Set for RA as a standardized reference for outcome assessment could inform the development of Core Outcome Sets (COS) for regulatory approval and improve comparability across studies. This would align RA research with broader international efforts to implement person-centered and functioning-oriented health systems. Policymakers, funding agencies, and regulatory bodies such as the FDA and EMA should consider mandating the inclusion of functioning measures in trial protocols, particularly for chronic conditions with major functional consequences. Such policy changes would promote more comprehensive, equitable, and patient-relevant evidence generation.

This review underscores the critical role of functioning as a core outcome in RA clinical trials and highlights the opportunity the ICF framework offers to enhance the relevance, comparability, and person-centeredness of pharmacological research. While functioning is often implicitly assessed, explicitly integrating the ICF Brief Core Set for RA would standardize outcome measurement and ensure that trials better reflect what matters most to patients. Bridging this gap between clinical efficacy and the lived experience of health is essential for advancing meaningful, policy-relevant, and patient-centered care in rheumatology.

## Conclusion

In summary, this review highlights that functioning is a critical yet under-recognized outcome in Phase III randomized clinical trials in rheumatoid arthritis. While many commonly used assessment tools implicitly reflect ICF categories, the explicit use of the ICF Brief Core Set for RA remains absent. Adopting the ICF as a standard framework for outcome measurements of RCTs would enhance the consistency, comparability, and patient-centeredness of outcome measurement in pharmacological research. Prioritizing functioning as a core outcome alongside clinical and biological markers is essential to ensure that RA trials capture what truly matters to patients and support more comprehensive treatment evaluations.

## Author Contributions

**AMDLT**: Conceptualization, Methodology, Supervision, Writing: Original Draft, Writing: Review & Editing. **PL**: Conceptualization, Methodology, Formal Analysis, Data Curation, Investigation, Writing: Original Draft, Visualization. **OA**: Conceptualization, Data Curation, Writing: Review & Editing. **RM**: Conceptualization, Methodology, Writing: Review & Editing, Supervision.

## Data Availability

All data supporting this review are contained within the article and supplementary materials.

## Ethics Approval

Not applicable.

## Declaration of generative AI and AI-assisted technologies in the writing process

During the preparation of this work the authors used ChatGPT 4.0 in order to find typographical errors. After using this tool/service, the authors reviewed and edited the content as needed and take full responsibility for the content of the published article.

## Acknowledgments.

We would like to thank Dr. Magda Gamba, medical librarian at ZHBL, for her input and insights for the search and extraction strategy.

