## Supplemental Material for "Use of the International Classification of Functioning, Disability and Health (ICF) in Randomized Controlled Trials of Rheumatoid Arthritis Pharmacological Treatments: a Scoping Review"

### (Supplement)

Adrian Martinez-De la Torre[a,b,d*#], Polina Leshetkina[a], Ogie Ahanor[c], Roxanne Maritz[a,d]

[a]*Faculty of Health Sciences and Medicine, University of Lucerne, Lucerne, Switzerland*

[b]*Swiss Paraplegic Research, Nottwil, Switzerland*

[c]*Private University in the Principality of Liechtenstein, Triesen, Lichtenstein*

[d] *Center for Rehabilitation in Global Health Systems, Faculty of Health Sciences and Medicine, University of Lucerne, Lucerne, Switzerland*

*Correspondence to: Adrian Martinez-De la Torre, Faculty of Health Sciences and Medicine, University of Lucerne, Lucerne, Switzerland. Frohburgstrasse, 6002, Lucerne, Switzerland;

[#] Both authors contributed equally to this work.

Adrian Martinez-De la Torre, ORCID: https://orcid.org/0000-0002-3155-7421
Roxanne Maritz, ORCID: https://orcid.org/0000-0002-5711-0388

***Table S1.*** *Preferred Reporting Items for Systematic reviews and Meta-Analyses extension for Scoping Reviews (PRISMA-ScR) Checklist.*

| SECTION | ITEM | PRISMA-ScR CHECKLIST ITEM | REPORTED ON PAGE # |
|---|---|---|---|
| **TITLE** | | | |
| Title | 1 | Identify the report as a scoping review. | Title Section |
| **ABSTRACT** | | | |
| Structured summary | 2 | Provide a structured summary that includes (as applicable): background, objectives, eligibility criteria, sources of evidence, charting methods, results, and conclusions that relate to the review questions and objectives. | Abstract |
| **INTRODUCTION** | | | |
| Rationale | 3 | Describe the rationale for the review in the context of what is already known. Explain why the review questions/objectives lend themselves to a scoping review approach. | Introduction, paragraphs 1-3 |
| Objectives | 4 | Provide an explicit statement of the questions and objectives being addressed with reference to their key elements (e.g., population or participants, concepts, and context) or other relevant key elements used to conceptualize the review questions and/or objectives. | Last paragraph introduction |
| **METHODS** | | | |
| Protocol and registration | 5 | Indicate whether a review protocol exists; state if and where it can be accessed (e.g., a Web address); and if available, provide registration information, including the registration number. | Methods: Protocol and Registration paragraph |
| Eligibility criteria | 6 | Specify characteristics of the sources of evidence used as eligibility criteria (e.g., years considered, language, and publication status), and provide a rationale. | Table 1 |

| | | | |
|---|---|---|---|
| Information sources | 7 | Describe all information sources in the search (e.g., databases with dates of coverage and contact with authors to identify additional sources), as well as the date the most recent search was executed. | Methods: information sources and search strategy |
| Search | 8 | Present the full electronic search strategy for at least 1 database, including any limits used, such that it could be repeated. | Table 2 |
| Selection of sources of evidence | 9 | State the process for selecting sources of evidence (i.e., screening and eligibility) included in the scoping review. | Methods: Study selection and data collection |
| Data charting process | 10 | Describe the methods of charting data from the included sources of evidence (e.g., calibrated forms or forms that have been tested by the team before their use, and whether data charting was done independently or in duplicate) and any processes for obtaining and confirming data from investigators. | Methods: Data extraction and critical appraisal |
| Data items | 11 | List and define all variables for which data were sought and any assumptions and simplifications made. | Methods: Data extraction and critical appraisal |
| Critical appraisal of individual sources of evidence | 12 | If done, provide a rationale for conducting a critical appraisal of included sources of evidence; describe the methods used and how this information was used in any data synthesis (if appropriate). | NA |
| Synthesis of results | 13 | Describe the methods of handling and summarizing the data that were charted. | Methods: Data analysis and presentation |
| **RESULTS** | | | |
| Selection of sources of evidence | 14 | Give numbers of sources of evidence screened, assessed for eligibility, and included in the review, with reasons for exclusions at each stage, ideally using a flow diagram. | Results: Study selection and Figure 1 |
| Characteristics of sources of | 15 | For each source of evidence, present characteristics for which data were charted and provide the citations. | Results: Characteristics of included studies + |

| | | | |
|---|---|---|---|
| evidence | | | Table S2 |
| Critical appraisal within sources of evidence | 16 | If done, present data on critical appraisal of included sources of evidence (see item 12). | NA |
| Results of individual sources of evidence | 17 | For each included source of evidence, present the relevant data that were charted that relate to the review questions and objectives. | Table 3-4 |
| Synthesis of results | 18 | Summarize and/or present the charting results as they relate to the review questions and objectives. | Results: Linking outcomes…+ Figure 2 |
| **DISCUSSION** | | | |
| Summary of evidence | 19 | Summarize the main results (including an overview of concepts, themes, and types of evidence available), link to the review questions and objectives, and consider the relevance to key groups. | Discussion: Paragraphs 1-4 |
| Limitations | 20 | Discuss the limitations of the scoping review process. | Discussion: Limitations |
| Conclusions | 21 | Provide a general interpretation of the results with respect to the review questions and objectives, as well as potential implications and/or next steps. | Discussion: Last 3 paragraphs + Conclusion |
| **FUNDING** | | | |
| Funding | 22 | Describe sources of funding for the included sources of evidence, as well as sources of funding for the scoping review. Describe the role of the funders of the scoping review. | Funding |

***Table S2.*** *Descriptive information of included studies.*

| Author and year | Title | Drug class | Country | Total sample size | Reference number |
|---|---|---|---|---|---|
| Liu 2025 | Ivarmacitinib, a selective Janus kinase 1 inhibitor, in patients with moderate-to-severe active rheumatoid arthritis and inadequate response to conventional synthetic DMARDs: results from a phase III randomized clinical trial. | SMARDs | multicentered | 566 | [1] |
| Van Vollenhoven 2024 | Upadacitinib monotherapy versus methotrexate monotherapy in patients with rheumatoid arthritis: efficacy and safety through 5 years in the SELECT-EARLY randomized controlled trial. | SMARDs and tDMARDs | Netherlands; US; Japan; Guatemala; Argentina; Poland | 945 | [2] |
| Smolen 2024 | Efficacy and safety of CT-P47 versus reference tocilizumab: 32-week results of a randomized, active-controlled, double-blind, phase III study in patients with rheumatoid arthritis, including 8 weeks of switching data from reference tocilizumab to CT-P47. | bDMARDs | Poland | 471 | [3] |
| Mazurov 2024 | Efficacy and Safety of Levilimab in Combination with Methotrexate in Patients with Active Rheumatoid Arthritis: 56-Week Results of Phase III Randomized Double-Blind Placebo-Controlled Trial SOLAR. | bDMARDs and tDMARDs | Russia; Belarus | 154 | [4] |
| Leng 2024 | A phase 3, randomized, double-blind, active-controlled clinical trial to compare BAT1806/BIIB800, a tocilizumab biosimilar, with tocilizumab reference product in participants with moderate-to-severe rheumatoid arthritis with inadequate response to methotr | bDMARDs | multicentered | 621 | [5] |
| Combe 2021 | Filgotinib versus placebo or adalimumab in patients with rheumatoid arthritis and inadequate response to methotrexate: a phase III randomised clinical trial. | SMARDs bDMARDs | multicentered | 1755 | [6] |
| Kameda 2020 | Sarilumab monotherapy or in combination with non-methotrexate disease-modifying antirheumatic drugs in active rheumatoid arthritis: A Japan phase 3 trial (HARUKA). | SMARDs | Japan | 91 | [7] |
| Genovese 2020 | Comparative clinical efficacy and safety of the proposed biosimilar ABP 710 with infliximab reference product in patients with rheumatoid arthritis. | bDMARDs | US; Spain; Hungary; Germany; Czech Republic; Poland | 558 | [8] |
| Cohen 2020 | Long-term Efficacy, Safety, and Immunogenicity of the Infliximab (IFX) Biosimilar, PF-06438179/GP1111, in Patients with Rheumatoid Arthritis After Switching from Reference IFX or Continuing Biosimilar Therapy: Week 54-78 Data From a Randomized, Double-Bli | bDMARDs | US; Japan; Brazil; Germany; Philippines; Korea; Canada; UK and Northern Ireland | 566 | [9] |
| Tanaka 2019 | Efficacy and safety of peficitinib (ASP015K) in patients with rheumatoid arthritis and an inadequate response to conventional DMARDs: a randomized, double-blind, placebo-controlled phase III trial (RAJ3). | SMARDs | Japan; Korea; Taiwan, Province of China | 507 | [10] |

| | | | | | |
|---|---|---|---|---|---|
| Suh 2019 | Long-Term Efficacy and Safety of Biosimilar CT-P10 Versus Innovator Rituximab in Rheumatoid Arthritis: 48-Week Results from a Randomized Phase III Trial. | Biologic DMARDs | Korea; Peru; Mexico; Chile; Poland; Germany; Colombia; Ukraine; Latvia | 372 | [11] |
| Lila 2019 | A phase III study of BCD-055 compared with innovator infliximab in patients with active rheumatoid arthritis: 54-week results from the LIRA study. | Biologic DMARDs | Russian Federation; Belarus; India | 426 | [12] |
| Tanaka 2019 | Sarilumab plus methotrexate in patients with active rheumatoid arthritis and inadequate response to methotrexate: results of a randomized, placebo-controlled phase III trial in Japan. | bDMARDs and tDMARDs | Japan | 243 | [13] |
| Rezaieyazdi 2019 | International multicenter randomized, placebo-controlled phase III clinical trial of beta-D-mannuronic acid in rheumatoid arthritis patients. | Experimental Agents | Iran; Pakistan | 288 | [14] |
| Tanaka 2019 | Modified- versus immediate-release tofacitinib in Japanese rheumatoid arthritis patients: a randomized, phase III, non-inferiority study. | SMARDs | Japan | 209 | [15] |
| Strand 2019 | Effects of upadacitinib on patient-reported outcomes: results from SELECT-BEYOND, a phase 3 randomized trial in patients with rheumatoid arthritis and inadequate responses to biologic disease-modifying antirheumatic drugs. | SMARDs | US; Germany; Canada | 498 | [16] |
| Strand 2019 | Patient-reported outcomes for tofacitinib with and without methotrexate, or adalimumab with methotrexate, in rheumatoid arthritis: a phase IIIB/IV trial. | SMARDs and bDMARDs | US; Argentina; UK and Northern Ireland; Austria | 1152 | [17] |
| Edwards 2019 | Safety of adalimumab biosimilar MSB11022 (acetate-buffered formulation) in patients with moderately-to-severely active rheumatoid arthritis. | bDMARDs | UK and Northern Ireland; Switzerland; US | 288 | [18] |
| Cohen 2018 | Similar efficacy, safety and immunogenicity of adalimumab biosimilar BI 695501 and Humira reference product in patients with moderately to severely active rheumatoid arthritis: results from the phase III randomized VOLTAIRE-RA equivalence study. | bDMARDs | US; Spain; Poland; Germany | 645 | [19] |
| Taylor 2018 | Efficacy and safety of monotherapy with sirukumab compared with adalimumab monotherapy in biologic-naive patients with active rheumatoid arthritis (SIRROUND-H): a randomized, double-blind, parallel-group, multinational, 52-week, phase 3 study. | bDMARDs | US, Europe, Latin America and South Africa | 559 | [20] |
| Matsuno 2018 | Phase III, multicenter, double-blind, randomized, parallel-group study to evaluate the similarities between LBEC0101 and etanercept reference product in terms of efficacy and safety in patients with active rheumatoid arthritis inadequately responding to m | bDMARDs | Japan; Korea | 374 | [21] |
| Smolen 2018 | Safety, immunogenicity and efficacy after switching from reference infliximab to biosimilar SB2 compared with continuing reference infliximab and SB2 in patients with | bDMARDs | Austria; Korea;US | 584 | [22] |

| | | | | | |
|---|---|---|---|---|---|
| | rheumatoid arthritis: results of a randomized, double-blind, phase III transition study. | | | | |
| Park 2018 | Comparison of biosimilar CT-P10 and innovator rituximab in patients with rheumatoid arthritis: a randomized controlled Phase 3 trial. | bDMARDs | Korea; US | 372 | [23] |
| Strand 2018 | Patient-reported outcomes from a randomized phase III trial of sarilumab monotherapy versus adalimumab monotherapy in patients with rheumatoid arthritis. | bDMARDs | US | 396 | [24] |
| Genovese 2018 | Two years of sarilumab in patients with rheumatoid arthritis and an inadequate response to MTX: safety, efficacy and radiographic outcomes | bDMARDs and tDMARDs | US | 901 | [25] |
| Takeuchi 2018 | Sirukumab in rheumatoid arthritis refractory to sulfasalazine or methotrexate: a randomized phase 3 safety and efficacy study in Japanese patients. | bDMARDs and tDMARDs | Japan | 122 | [26] |
| Weinblatt 2018 | Switching From Reference Adalimumab to SB5 (Adalimumab Biosimilar) in Patients With Rheumatoid Arthritis: Fifty-Two-Week Phase III Randomized Study Results. | bDMARDs and tDMARDs | US;Lithuania; Czech Republic; Poland; Korea; Bosnia and Herzegovina; Bulgaria; Ukraine | 544 | [27] |
| Ogata 2018 | A randomized, double-blind, parallel-group, phase III study of shortening the dosing interval of subcutaneous tocilizumab monotherapy in patients with rheumatoid arthritis and an inadequate response to subcutaneous tocilizumab every other week: Results of | bDMARDs | Japan | 340 | [28] |
| Takeuchi 2017 | Sirukumab for rheumatoid arthritis: the phase III SIRROUND-D study. | bDMARDs | UK and Northern Ireland | 1670 | [29] |
| Cohen 2017 | Efficacy and safety of the biosimilar ABP 501 compared with adalimumab in patients with moderate to severe rheumatoid arthritis: a randomised, double-blind, phase III equivalence study. | bDMARDs | US; UK and Northern Ireland; Spain; Germany; Czech Republic; Poland | 526 | [30] |
| Burmester 2017 | Efficacy and safety of sarilumab monotherapy versus adalimumab monotherapy for the treatment of patients with active rheumatoid arthritis (MONARCH): a randomised, double-blind, parallel-group phase III trial. | bDMARDs | Germany;US; Chile; Korea | 369 | [31] |
| Dougados 2017 | Baricitinib in patients with inadequate response or intolerance to conventional synthetic DMARDs: results from the RA-BUILD study. | SMARDs | France; Netherlands; Taiwan, Province of China;US; Hungary; UK and Northern Ireland | 684 | [32] |
| Bae 2017 | A phase III, multicenter, randomized, double-blind, active-controlled, parallel-group trial comparing safety and efficacy of HD203, with innovator etanercept, in combination with methotrexate, in patients with rheumatoid arthritis: the HERA study. | bDMARDs and tDMARDs | Korea | 294 | [33] |

| | | | | | |
|---|---|---|---|---|---|
| Emery 2017 | A phase III randomised, double-blind, parallel-group study comparing SB4 with etanercept reference product in patients with active rheumatoid arthritis despite methotrexate therapy. | bDMARDs and tDMARDs | UK and Northern Ireland | 596 | [34] |
| Jamshidi 2017 | A phase III, randomized, two-armed, double-blind, parallel, active controlled, and non-inferiority clinical trial to compare efficacy and safety of biosimilar adalimumab (CinnoRA R) to the reference product (Humira R) in patients with active rheumatoid ar | bDMARDs | Iran | 136 | [35] |
| Schiff 2017 | Patient-reported outcomes of baricitinib in patients with rheumatoid arthritis and no or limited prior disease-modifying antirheumatic drug treatment. | SMARDs | US | 588 | [82] |
| Taylor 2017 | Baricitinib versus Placebo or Adalimumab in Rheumatoid Arthritis. | SMARDs and bDMARDs | multicentered | 1307 | [37] |
| Fleischmann 2017 | Sarilumab and Nonbiologic Disease-Modifying Antirheumatic Drugs in Patients With Active Rheumatoid Arthritis and Inadequate Response or Intolerance to Tumor Necrosis Factor Inhibitors. | bDMARDs and tDMARDs. | US; Australia; Canada; Czech Republic; Germany; Greece; Hungary; Israel; Italy | 546 | [38] |
| Fleischmann 2017 | Baricitinib, Methotrexate, or Combination in Patients With Rheumatoid Arthritis and No or Limited Prior Disease-Modifying Antirheumatic Drug Treatment. | SMARDs and tDMARDs | US; Colombia; Netherlands; Mexico; Argentina; Russia; Brazil; India; Japan | 588 | [39] |
| Strand 2017 | Tofacitinib in Combination With Conventional Disease-Modifying Antirheumatic Drugs in Patients With Active Rheumatoid Arthritis: Patient-Reported Outcomes From a Phase III Randomized Controlled Trial. | SMARDs and tDMARDs | US | 795 | [40] |
| Burmester 2016 | Tocilizumab in early progressive rheumatoid arthritis: FUNCTION, a randomized controlled trial. | bDMARDs | Germany | 1157 | [41] |
| Strand 2016 | Sarilumab plus methotrexate improves patient-reported outcomes in patients with active rheumatoid arthritis and inadequate responses to methotrexate: results of a phase III trial. | bDMARDs and tDMARDs | US | 1197 | [42] |
| Yoo 2016 | A phase III randomized study to evaluate the efficacy and safety of CT-P13 compared with reference infliximab in patients with active rheumatoid arthritis: 54-week results from the PLANETRA study. | Experimental agents and bDMARDs | Korea; US | 606 | [43] |
| Genovese 2016 | Baricitinib in Patients with Refractory Rheumatoid Arthritis. | bDMARDs | multicentered | 527 | [44] |
| Strand 2016 | Tofacitinib or adalimumab versus placebo: patient-reported outcomes from a phase 3 study of active rheumatoid arthritis. | SMARDs and bDMARDs | US; Canada; Sweden; Korea | 717 | [78] |
| Fleischmann 2016 | Patient-Reported Outcomes From a Two-Year Head-to-Head Comparison of Subcutaneous Abatacept and Adalimumab for Rheumatoid Arthritis. | bDMARDs | US | 646 | [46] |

| | | | | | |
|---|---|---|---|---|---|
| Smolen 2015 | Certolizumab pegol in rheumatoid arthritis patients with low to moderate activity: the CERTAIN double-blind, randomized, placebo-controlled trial. | bDMARDs | Austria; France; Germany; Italy; Poland | 194 | [47] |
| Genovese 2015 | Sarilumab Plus Methotrexate in Patients With Active Rheumatoid Arthritis and Inadequate Response to Methotrexate: Results of a Phase III Study. | bDMARDs and tDMARDs | multicentered | 1369 | [48] |
| Choi 2014 | Comparison of the efficacy and safety profiles of a pelubiprofen versus celecoxib in patients with rheumatoid arthritis: a 6-week, multicenter, randomized, double-blind, phase III, non-inferiority clinical trial. | NSAIDs | Korea | 145 | [49] |
| Yamamoto 2014 | Efficacy and safety of certolizumab pegol without methotrexate co-administration in Japanese patients with active rheumatoid arthritis: the HIKARI randomized, placebo-controlled trial. | bDMARDs | Japan | 230 | [50] |
| Bingham 2014 | The effect of intravenous golimumab on health-related quality of life in rheumatoid arthritis: 24-week results of the phase III GO-FURTHER trial. | bDMARDs | US | 592 | [51] |
| Ogata 2014 | Phase III study of the efficacy and safety of subcutaneous versus intravenous tocilizumab monotherapy in patients with rheumatoid arthritis. | bDMARDs | Japan | 348 | [52] |
| Yoo 2013 | A randomised, double-blind, parallel-group study to demonstrate equivalence in efficacy and safety of CT-P13 compared with innovator infliximab when coadministered with methotrexate in patients with active rheumatoid arthritis: the PLANETRA study. | bDMARDs and tDMARDs | Korea; Poland; Chile; Philippines; Ukraine; Bosnia and Herzegovina; Colombia; Mexico; Peru;US; Austria; Germany; Italy; Jordan | 606 | [53] |
| Takeuchi 2013 | Golimumab monotherapy in Japanese patients with active rheumatoid arthritis despite prior treatment with disease-modifying antirheumatic drugs: results of the phase 2/3, multicentre, randomised, double-blind, placebo-controlled GO-MONO study through 24 we | bDMARDs | Japan | 316 | [54] |
| Kim 2013 | A clinical trial and extension study of infliximab in Korean patients with active rheumatoid arthritis despite methotrexate treatment. | bDMARDs and tDMARDs | Korea | 143 | [55] |
| Stohl 2012 | Safety and efficacy of ocrelizumab in combination with methotrexate in MTX-naive subjects with rheumatoid arthritis: the phase III FILM trial. | bDMARDs and tDMARDs | US; UK and Northern Ireland; Spain; Switzerland; Brazil | 613 | [56] |
| Yazici 2012 | Efficacy of tocilizumab in patients with moderate to severe active rheumatoid arthritis and a previous inadequate response to disease-modifying antirheumatic drugs: the ROSE study. | bDMARDs | US | 619 | [57] |

| | | | | | |
|---|---|---|---|---|---|
| Fleischmann 2012 | Placebo-controlled trial of tofacitinib monotherapy in rheumatoid arthritis. | SMARDs | multicentered | 611 | [58] |
| Strand 2012 | Health-related quality of life outcomes of adalimumab for patients with early rheumatoid arthritis: results from a randomized multicenter study. | bDMARDs | US | 799 | [59] |
| Taylor 2011 | Ofatumumab, a fully human anti-CD20 monoclonal antibody, in biological-naive, rheumatoid arthritis patients with an inadequate response to methotrexate: a randomized, double-blind, placebo-controlled clinical trial. | bDMARDs and tDMARDs | UK and Northern Ireland;US; Denmark | 265 | [60] |
| Emery 2010 | Efficacy and safety of different doses and retreatment of rituximab: a randomized, placebo-controlled trial in patients who are biological naive with active rheumatoid arthritis and an inadequate response to methotrexate (Study Evaluating Rituximab's Effi | bDMARDs and tDMARDs | UK and and Northern Ireland;US; France; Poland; Mexico | 512 | [61] |
| Genovese 2011 | Subcutaneous abatacept versus intravenous abatacept: a phase IIIb noninferiority study in patients with an inadequate response to methotrexate. | bDMARDs and tDMARDs | US; Mexico; Peru; Argentina; Brazil; Australia; Poland; Russia; Belgium; Germany | 1457 | [62] |
| Rubbert-Roth 2010 | Efficacy and safety of various repeat treatment dosing regimens of rituximab in patients with active rheumatoid arthritis: results of a Phase III randomized study (MIRROR). | bDMARDs | Germany; Netherlands; Brazil; Canada; Spain; UK and Northern Ireland | 378 | [63] |
| Yang 2024 | Safety and efficacy of peficitinib in Asian patients with rheumatoid arthritis who had an inadequate response or intolerance to methotrexate: results of a multicenter, randomized, double-blind, placebo-controlled phase 3 study | SMARDs and tDMARDs | China, Korea, and Taiwan | 385 | [64] |
| Taylor 2023 | Anti-GM-CSF otilimab versus sarilumab or placebo in patients with rheumatoid arthritis and inadequate response to targeted therapies: a phase III randomized trial (contRAst 3) | bDMARDs | UK and Northern Ireland;US; Japan; Ireland | 549 | [65] |
| Tanaka 2023 | Efficacy and safety of the anti-TNF multivalent NANOBODY¬Æ compound ozoralizumab in patients with rheumatoid arthritis and an inadequate response to methotrexate: A 52-week result of a Phase II/III study (OHZORA trial) | bDMARDs | multicentered | 381 | [66] |
| Ye 2023 | Efficacy and Safety of CMAB008 Compared with Innovator Infliximab in Patients with Moderate-to-Severe Rheumatoid Arthritis Receiving Concomitant Methotrexate: A Randomized, Double-blind, Multi-center, Phase III Non-inferiority Study | bDMARDs and tDMARDs | China | 384 | [67] |
| Curtis 2023 | Effects of Disease-Worsening Following Withdrawal of Etanercept or Methotrexate on Patient-Reported Outcomes in Patients With Rheumatoid Arthritis: Results From the SEAM-RA Trial | bDMARDs and tDMARDs | US; Canada; UK and Northern Ireland | 253 | [68] |

| | | | | | |
|---|---|---|---|---|---|
| Takeuchi 2022 | Phase II/III Results of a Trial of Anti‚ÄìTumor Necrosis Factor Multivalent NANOBODY Compound Ozoralizumab in Patients With Rheumatoid Arthritis | bDMARDs | multicentred | 395 | [69] |
| Furst 2022 | Efficacy and safety of switching from reference adalimumab to CT-P17 (100 mg/ml): 52-week randomized, double-blind study in rheumatoid arthritis | bDMARDs | multicentred | 607 | [70] |
| Liu 2022 | Fine Comparison of the Efficacy and Safety Between GB242 and Infliximab in Patients with Rheumatoid Arthritis: A Phase III Study | bDMARDs | China | 570 | [71] |
| Nasonov 2022 | Olokizumab, a monoclonal antibody against interleukin 6, in combination with methotrexate in patients with rheumatoid arthritis inadequately controlled by methotrexate: efficacy and safety results of a randomized controlled phase III study | bDMARDs and tDMARDs | Russia; Belarus; Bulgaria; US | 428 | [72] |
| Kay 2021 | Efficacy and safety of biosimilar CT-P17 versus reference adalimumab in subjects with rheumatoid arthritis: 24-week results from a randomized study | bDMARDs | Canada; Bulgaria; Hungary; Lithuania; Peru; Poland; Ukraine;US; Italy | 648 | [73] |
| Behrens 2021 | Rituximab plus leflunomide in rheumatoid arthritis: A randomized, placebo-controlled, investigator-initiated clinical trial (AMARA study) | bDMARDs and tDMARDs | Germany; Switzerland; UK and Northern Ireland | 140 | [74] |
| Westhovens 2021 | Filgotinib in combination with methotrexate or as monotherapy versus methotrexate monotherapy in patients with active rheumatoid arthritis and limited or no prior exposure to methotrexate: The phase 3, randomized controlled FINCH 3 trial | SMARDs and tDMARDs | Belgium; US; Netherlands; New Zealand; Canada; India; Argentina; Germany; Japan | 1252 | [75] |
| Wiland 2020 | Switching to Biosimilar SDZ-ADL in Patients with Moderate-to-Severe Active Rheumatoid Arthritis: 48-Week Efficacy, Safety and Immunogenicity Results From the Phase III, Randomized, Double-Blind ADMYRA Study | bDMARDs | Poland; US; Germany; Spain; Czech Republic; UK and Northern Ireland | 353 | [76] |
| Yamanaka 2020 | A Comparative Study to Assess the Efficacy, Safety, and Immunogenicity of YLB113 and the Etanercept Reference Product for the Treatment of Patients with Rheumatoid Arthritis | bDMARDs | Europe, Japan; India | 528 | [77] |
| Alten 2019 | Randomised, double-blind, phase III study comparing the infliximab biosimilar, PF-06438179/GP1111, with reference infliximab: Efficacy, safety and immunogenicity from week 30 to week 54 | bDMARDs | Germany; Poland; Czech Republic; Japan; Brazil; Ukraine; US; UK and Northern Ireland | 650 | [78] |
| Matucci-Cerinic 2018 | Efficacy, safety and immunogenicity of GP2015, an etanercept biosimilar, compared with the reference etanercept in patients with moderate-To-severe rheumatoid arthritis: 24-week results from the comparative phase III, randomised, double-blind EQUIRA study | bDMARDs | Italy; France; US; Germany; Poland | 376 | [79] |

| | | | | | |
|---|---|---|---|---|---|
| Smolen 2017 | Patient-reported outcomes from a randomised phase III study of baricitinib in patients with rheumatoid arthritis and an inadequate response to biological agents (RA-BEACON) | SMARDs | Austria; US; Australia; Spain; France | 527 | [80] |
| Chen 2016 | A randomized, controlled trial of efficacy and safety of Anbainuo, a bio-similar etanercept, for moderate to severe rheumatoid arthritis inadequately responding to methotrexate | SMARDs | China | 600 | [81] |
| Takeuchi 2015 | Evaluation of the pharmacokinetic equivalence and 54-week efficacy and safety of CT-P13 and innovator infliximab in Japanese patients with rheumatoid arthritis | bDMARDs | Japan; Korea | 104 | [82] |
| Smolen 2015 | Efficacy and safety of tabalumab, an anti-B-cellactivating factor monoclonal antibody, in patients with rheumatoid arthritis who had an inadequate response to methotrexate therapy: Results from a phase III multicentre, randomised, double-blind study | bDMARDs | multicentered | 1041 | [83] |
| Genovese 2014 | A phase III, multicenter, randomized, double-blind, placebo-controlled, parallel-group study of 2 dosing regimens of fostamatinib in patients with rheumatoid arthritis with an inadequate response to a tumor necrosis factor-Œ± antagonist | Other | US; Netherlands; Canada; Argentina; France; UK and Northern Ireland;Australia | 323 | [84] |
| Takeuchi 2014 | Adalimumab, a human anti-TNF monoclonal antibody, outcome study for the prevention of joint damage in Japanese patients with early rheumatoid arthritis: The HOPEFUL 1 study | bDMARDs | multicentered | 171 | [85] |
| Genovese 2012 | Effect of golimumab on patient-reported outcomes in rheumatoid arthritis: Results from the GO-FORWARD study | bDMARDs | US; Canada | 444 | [86] |
| Kameda2020 | A Phase 2b/3, Randomized, Double-Blind Study Comparing Upadacitinib (ABT-494) to Placebo in Japanese Subjects With Moderately to Severely Active Rheumatoid Arthritis Who Are on a Stable Dose of Conventional Synthetic Disease-Modifying Anti-Rheumatic Drugs | SMARDs and tDMARDs. | Japan | 197 | [87] |
| Park2018 | A Randomized, Controlled, Double-Blind, Parallel-Group, Phase 3 Study to Compare the Pharmacokinetics, Efficacy and Safety Between CT-P10, Rituxan and MabThera in Patients With Rheumatoid Arthritis | bDMARDs | Europe, Asia Pacific, and Latin America | 372 | [88] |
| Fleischmann2017 | A Randomized, Double-blind, Parallel, Placebo-controlled Study Assessing the Efficacy and Safety of Sarilumab Added to Non-biologic DMARD Therapy in Patients With Rheumatoid Arthritis Who Are Inadequate Responders to or Intolerant of TNF-Œ± Antagonists | bDMARDs and tDMARDs | multicenter | 546 | [89] |
| Genovese 2020 | Efficacy and safety of sarilumab in combination with csDMARDs or as monotherapy in subpopulations of patients with moderately to severely active rheumatoid arthritis in three phase III randomized, controlled studies | bDMARDs | US; UK and Northern Ireland; Germany; Korea | 2108 | [90] |
| Coombs 2010 | Improved pain, physical functioning and health status in patients with rheumatoid arthritis treated with CP-690,550, | SMARDs | US; Netherlands; Mexico; Brazil | 264 | [91] |

| | |
|---|---|
| | an orally active Janus kinase (JAK) inhibitor: results from a randomized, double-blind, placebo-controlled trial. |

9 Cohen SB, Radominski SC, Kameda H, *et al.* Long-term Efficacy, Safety, and Immunogenicity of the Infliximab (IFX) Biosimilar, PF-06438179/GP1111, in Patients with Rheumatoid Arthritis After Switching from Reference IFX or Continuing Biosimilar Therapy: Week 54-78 Data From a Randomized, Double-Blind, Phase III Trial.

*BioDrugs : clinical immunotherapeutics, biopharmaceuticals and gene therapy*. 2020;34. doi: 10.1007/s40259-019-00403-z

10 Tanaka Y, Takeuchi T, Tanaka S, *et al.* Efficacy and safety of peficitinib (ASP015K) in patients with rheumatoid arthritis and an inadequate response to conventional DMARDs: a randomised, double-blind, placebo-controlled phase III trial (RAJ3). *Annals of the rheumatic diseases*. 2019;78. doi: 10.1136/annrheumdis-2019-215163

11 Suh C-H, Yoo DH, Berrocal Kasay A, *et al.* Long-Term Efficacy and Safety of Biosimilar CT-P10 Versus Innovator Rituximab in Rheumatoid Arthritis: 48-Week Results from a Randomized Phase III Trial. *BioDrugs : clinical immunotherapeutics, biopharmaceuticals and gene therapy*. 2019;33. doi: 10.1007/s40259-018-00331-4

12 Lila AM, Mazurov VI, Denisov LN, *et al.* A phase III study of BCD-055 compared with innovator infliximab in patients with active rheumatoid arthritis: 54-week results from the LIRA study. *Rheumatology international*. 2019;39. doi: 10.1007/s00296-019-04359-9

13 Tanaka Y, Wada K, Takahashi Y, *et al.* Sarilumab plus methotrexate in patients with active rheumatoid arthritis and inadequate response to methotrexate: results of a randomized, placebo-controlled phase III trial in Japan. *Arthritis research & therapy*. 2019;21. doi: 10.1186/s13075-019-1856-4

14 Rezaieyazdi Z, Farooqi A, Soleymani-Salehabadi H, *et al.* International multicenter randomized, placebo-controlled phase III clinical trial of beta-D-mannuronic acid in rheumatoid arthritis patients. *Inflammopharmacology*. 2019;27. doi: 10.1007/s10787-018-00557-2

15 Tanaka Y, Sugiyama N, Toyoizumi S, *et al.* Modified- versus immediate-release tofacitinib in Japanese rheumatoid arthritis patients: a randomized, phase III, non-inferiority study. *Rheumatology (Oxford, England)*. 2019;58. doi: 10.1093/rheumatology/key250

16 Strand V, Schiff M, Tundia N, *et al.* Effects of upadacitinib on patient-reported outcomes: results from SELECT-BEYOND, a phase 3 randomized trial in patients with rheumatoid arthritis and inadequate responses to biologic disease-modifying antirheumatic drugs. *Arthritis research & therapy*. 2019;21. doi: 10.1186/s13075-019-2059-8

17 Strand V, Mysler E, Moots RJ, *et al.* Patient-reported outcomes for tofacitinib with and without methotrexate, or adalimumab with methotrexate, in rheumatoid arthritis: a phase IIIB/IV trial. *RMD open*. 2019;5. doi: 10.1136/rmdopen-2019-001040

18 Edwards CJ, Monnet J, Ullmann M, *et al.* Safety of adalimumab biosimilar MSB11022 (acetate-buffered formulation) in patients with moderately-to-severely active rheumatoid arthritis. *Clinical rheumatology*. 2019;38. doi: 10.1007/s10067-019-04679-y

19 Cohen SB, Alonso-Ruiz A, Klimiuk PA, *et al.* Similar efficacy, safety and immunogenicity of adalimumab biosimilar BI 695501 and Humira reference product in patients with moderately to severely active rheumatoid arthritis: results from the phase III randomised VOLTAIRE-RA equivalence study. *Annals of the rheumatic diseases*. 2018;77. doi: 10.1136/annrheumdis-2017-212245

20 Taylor PC, Schiff MH, Wang Q, *et al.* Efficacy and safety of monotherapy with sirukumab compared with adalimumab monotherapy in biologic-naive patients with active rheumatoid arthritis (SIRROUND-H): a randomised, double-blind, parallel-group, multinational, 52-week, phase 3 study. *Annals of the rheumatic diseases*. 2018;77. doi: 10.1136/annrheumdis-2017-212496

21 Matsuno H, Tomomitsu M, Hagino A, *et al.* Phase III, multicentre, double-blind, randomised, parallel-group study to evaluate the similarities between LBEC0101 and etanercept reference product in terms of efficacy and safety in patients with active rheumatoid arthritis inadequately responding to methotrexate. *Annals of the rheumatic diseases*. 2018;77. doi: 10.1136/annrheumdis-2017-212172

22 Smolen JS, Choe J-Y, Prodanovic N, *et al.* Safety, immunogenicity and efficacy after switching from reference infliximab to biosimilar SB2 compared with continuing reference infliximab and SB2 in patients with rheumatoid arthritis: results of a randomised, double-blind, phase III transition study. *Annals of the rheumatic diseases*. 2018;77. doi: 10.1136/annrheumdis-2017-211741

23 Park W, Bozic-Majstorovic L, Milakovic D, *et al.* Comparison of biosimilar CT-P10 and innovator rituximab in patients with rheumatoid arthritis: a randomized controlled Phase 3 trial. *mAbs*. 2018;10. doi: 10.1080/19420862.2018.1487912

24 Strand V, Gossec L, Proudfoot CWJ, *et al.* Patient-reported outcomes from a randomized phase III trial of sarilumab monotherapy versus adalimumab monotherapy in patients with rheumatoid arthritis. *Arthritis research & therapy*. 2018;20. doi: 10.1186/s13075-018-1614-z

25 Genovese MC, van Adelsberg J, Fan C, *et al.* Two years of sarilumab in patients with rheumatoid arthritis and an inadequate response to MTX: safety, efficacy and radiographic outcomes. *Rheumatology (Oxford)*. 2018;57:1423–31. doi: 10.1093/rheumatology/key121

26 Takeuchi T, Yamanaka H, Harigai M, *et al.* Sirukumab in rheumatoid arthritis refractory to sulfasalazine or methotrexate: a randomized phase 3 safety and efficacy study in Japanese patients. *Arthritis research & therapy*. 2018;20. doi: 10.1186/s13075-018-1536-9

27 Weinblatt ME, Baranauskaite A, Dokoupilova E, *et al.* Switching From Reference Adalimumab to SB5 (Adalimumab Biosimilar) in Patients With Rheumatoid Arthritis: Fifty-Two-Week Phase III Randomized Study Results. *Arthritis & rheumatology (Hoboken, NJ)*. 2018;70. doi: 10.1002/art.40444

28 Ogata A, Tanaka Y, Ishii T, *et al.* A randomized, double-blind, parallel-group, phase III study of shortening the dosing interval of subcutaneous tocilizumab monotherapy in patients with rheumatoid arthritis and an inadequate response to subcutaneous tocilizumab every other week: Results of the 12-week double-blind period. *Modern rheumatology*. 2018;28. doi: 10.1080/14397595.2017.1332507

29 Takeuchi T, Thorne C, Karpouzas G, *et al.* Sirukumab for rheumatoid arthritis: the phase III SIRROUND-D study. *Annals of the rheumatic diseases*. 2017;76. doi: 10.1136/annrheumdis-2017-211328

30 Cohen S, Genovese MC, Choy E, *et al.* Efficacy and safety of the biosimilar ABP 501 compared with adalimumab in patients with moderate to severe rheumatoid arthritis: a randomised, double-blind, phase III equivalence study. *Annals of the rheumatic diseases*. 2017;76. doi: 10.1136/annrheumdis-2016-210459

31 Burmester GR, Lin Y, Patel R, *et al.* Efficacy and safety of sarilumab monotherapy versus adalimumab monotherapy for the treatment of patients with active rheumatoid arthritis (MONARCH): a randomised, double-blind, parallel-group phase III trial. *Annals of the rheumatic diseases*. 2017;76. doi: 10.1136/annrheumdis-2016-210310

32 Dougados M, van der Heijde D, Chen Y-C, *et al.* Baricitinib in patients with inadequate response or intolerance to conventional synthetic DMARDs: results from the RA-BUILD study. *Annals of the rheumatic diseases*. 2017;76. doi: 10.1136/annrheumdis-2016-210094

33 Bae S-C, Kim J, Choe J-Y, *et al.* A phase III, multicentre, randomised, double-blind, active-controlled, parallel-group trial comparing safety and efficacy of HD203, with innovator etanercept, in combination with methotrexate, in patients with rheumatoid arthritis: the HERA study. *Annals of the rheumatic diseases*. 2017;76. doi: 10.1136/annrheumdis-2015-207613

34 Emery P, Vencovsky J, Sylwestrzak A, *et al.* A phase III randomised, double-blind, parallel-group study comparing SB4 with etanercept reference product in patients with active rheumatoid arthritis despite methotrexate therapy. *Annals of the rheumatic diseases*. 2017;76. doi: 10.1136/annrheumdis-2015-207588

35 Jamshidi A, Gharibdoost F, Vojdanian M, *et al.* A phase III, randomized, two-armed, double-blind, parallel, active controlled, and non-inferiority clinical trial to compare efficacy and safety of biosimilar adalimumab (CinnoRA R) to the reference product (Humira R) in patients with active rheumatoid arthritis. *Arthritis research & therapy*. 2017;19. doi: 10.1186/s13075-017-1371-4

36 Schiff M, Takeuchi T, Fleischmann R, *et al.* Patient-reported outcomes of baricitinib in patients with rheumatoid arthritis and no or limited prior disease-modifying antirheumatic drug treatment. *Arthritis research & therapy*. 2017;19. doi: 10.1186/s13075-017-1410-1

37 Taylor PC, Keystone EC, van der Heijde D, *et al.* Baricitinib versus Placebo or Adalimumab in Rheumatoid Arthritis. *The New England journal of medicine*. 2017;376. doi: 10.1056/NEJMoa1608345

38 Fleischmann R, van Adelsberg J, Lin Y, *et al.* Sarilumab and Nonbiologic Disease-Modifying Antirheumatic Drugs in Patients With Active Rheumatoid Arthritis and Inadequate Response or Intolerance to Tumor Necrosis Factor Inhibitors. *Arthritis & rheumatology (Hoboken, NJ)*. 2017;69. doi: 10.1002/art.39944

39 Fleischmann R, Schiff M, van der Heijde D, *et al.* Baricitinib, Methotrexate, or Combination in Patients With Rheumatoid Arthritis and No or Limited Prior Disease-Modifying Antirheumatic Drug Treatment. *Arthritis & rheumatology (Hoboken, NJ)*. 2017;69. doi: 10.1002/art.39953

40 Strand V, Kremer JM, Gruben D, *et al.* Tofacitinib in Combination With Conventional Disease-Modifying Antirheumatic Drugs in Patients With Active Rheumatoid Arthritis: Patient-Reported Outcomes From a Phase III Randomized Controlled Trial. *Arthritis care & research*. 2017;69. doi: 10.1002/acr.23004

41 Burmester GR, Rigby WF, van Vollenhoven RF, *et al.* Tocilizumab in early progressive rheumatoid arthritis: FUNCTION, a randomised controlled trial. *Annals of the rheumatic diseases*. 2016;75. doi: 10.1136/annrheumdis-2015-207628

42 Strand V, Kosinski M, Chen C-I, *et al.* Sarilumab plus methotrexate improves patient-reported outcomes in patients with active rheumatoid arthritis and inadequate responses to methotrexate: results of a phase III trial. *Arthritis research & therapy*. 2016;18. doi: 10.1186/s13075-016-1096-9

43 Yoo DH, Racewicz A, Brzezicki J, *et al.* A phase III randomized study to evaluate the efficacy and safety of CT-P13 compared with reference infliximab in patients with active rheumatoid arthritis: 54-week results from the PLANETRA study. *Arthritis Res Ther*. 2016;18:82. doi: 10.1186/s13075-016-0981-6

44 Genovese MC, Kremer J, Zamani O, *et al.* Baricitinib in Patients with Refractory Rheumatoid Arthritis. *The New England journal of medicine*. 2016;374. doi: 10.1056/NEJMoa1507247

45 Strand V, van Vollenhoven RF, Lee EB, *et al.* Tofacitinib or adalimumab versus placebo: patient-reported outcomes from a phase 3 study of active rheumatoid arthritis. *Rheumatology (Oxford, England)*. 2016;55. doi: 10.1093/rheumatology/kev442

46 Fleischmann R, Weinblatt ME, Schiff M, *et al.* Patient-Reported Outcomes From a Two-Year Head-to-Head Comparison of Subcutaneous Abatacept and Adalimumab for Rheumatoid Arthritis. *Arthritis care & research*. 2016;68. doi: 10.1002/acr.22763

47 Smolen JS, Emery P, Ferraccioli GF, *et al.* Certolizumab pegol in rheumatoid arthritis patients with low to moderate activity: the CERTAIN double-blind, randomised, placebo-controlled trial. *Annals of the rheumatic diseases*. 2015;74. doi: 10.1136/annrheumdis-2013-204632

48 Genovese MC, Fleischmann R, Kivitz AJ, *et al.* Sarilumab Plus Methotrexate in Patients With Active Rheumatoid Arthritis and Inadequate Response to Methotrexate: Results of a Phase III Study. *Arthritis & rheumatology (Hoboken, NJ)*. 2015;67. doi: 10.1002/art.39093

49 Choi IA, Baek H-J, Cho C-S, *et al.* Comparison of the efficacy and safety profiles of a pelubiprofen versus celecoxib in patients with rheumatoid arthritis: a 6-week, multicenter, randomized, double-blind, phase III, non-inferiority clinical trial. *BMC musculoskeletal disorders*. 2014;15. doi: 10.1186/1471-2474-15-375

50 Yamamoto K, Takeuchi T, Yamanaka H, *et al.* Efficacy and safety of certolizumab pegol without methotrexate co-administration in Japanese patients with active rheumatoid arthritis: the HIKARI randomized, placebo-controlled trial. *Modern rheumatology*. 2014;24. doi: 10.3109/14397595.2013.843764

51 Bingham CO 3rd, Weinblatt M, Han C, *et al.* The effect of intravenous golimumab on health-related quality of life in rheumatoid arthritis: 24-week results of the phase III GO-FURTHER trial. *The Journal of rheumatology*. 2014;41. doi: 10.3899/jrheum.130864

52 Ogata A, Tanimura K, Sugimoto T, *et al.* Phase III study of the efficacy and safety of subcutaneous versus intravenous tocilizumab monotherapy in patients with rheumatoid arthritis. *Arthritis care & research*. 2014;66. doi: 10.1002/acr.22110

53 Yoo DH, Hrycaj P, Miranda P, *et al.* A randomised, double-blind, parallel-group study to demonstrate equivalence in efficacy and safety of CT-P13 compared with innovator infliximab when coadministered with methotrexate in patients with active rheumatoid arthritis: the PLANETRA study. *Annals of the rheumatic diseases*. 2013;72. doi: 10.1136/annrheumdis-2012-203090

54 Takeuchi T, Harigai M, Tanaka Y, *et al.* Golimumab monotherapy in Japanese patients with active rheumatoid arthritis despite prior treatment with disease-modifying antirheumatic drugs: results of the phase 2/3, multicentre, randomised, double-blind, placebo-controlled GO-MONO study through 24 weeks. *Annals of the rheumatic diseases*. 2013;72. doi: 10.1136/annrheumdis-2012-201796

55 Kim J, Ryu H, Yoo D-H, *et al.* A clinical trial and extension study of infliximab in Korean patients with active rheumatoid arthritis despite methotrexate treatment. *Journal of Korean medical science*. 2013;28. doi: 10.3346/jkms.2013.28.12.1716

56 Stohl W, Gomez-Reino J, Olech E, *et al.* Safety and efficacy of ocrelizumab in combination with methotrexate in MTX-naive subjects with rheumatoid arthritis: the phase III FILM trial. *Annals of the rheumatic diseases*. 2012;71. doi: 10.1136/annrheumdis-2011-200706

57 Yazici Y, Curtis JR, Ince A, *et al.* Efficacy of tocilizumab in patients with moderate to severe active rheumatoid arthritis and a previous inadequate response to disease-modifying antirheumatic drugs: the ROSE study. *Annals of the rheumatic diseases*. 2012;71. doi: 10.1136/ard.2010.148700

58 Fleischmann R, Kremer J, Cush J, *et al.* Placebo-controlled trial of tofacitinib monotherapy in rheumatoid arthritis. *The New England journal of medicine*. 2012;367. doi: 10.1056/NEJMoa1109071

59 Strand V, Rentz AM, Cifaldi MA, *et al.* Health-related quality of life outcomes of adalimumab for patients with early rheumatoid arthritis: results from a randomized multicenter study. *The Journal of rheumatology*. 2012;39. doi: 10.3899/jrheum.101161

60 Taylor PC, Quattrocchi E, Mallett S, *et al.* Ofatumumab, a fully human anti-CD20 monoclonal antibody, in biological-naive, rheumatoid arthritis patients with an inadequate response to methotrexate: a randomised, double-blind, placebo-controlled clinical trial. *Annals of the rheumatic diseases*. 2011;70. doi: 10.1136/ard.2011.151522

61 Emery P, Deodhar A, Rigby WF, *et al.* Efficacy and safety of different doses and retreatment of rituximab: a randomised, placebo-controlled trial in patients who are biological naive with active rheumatoid arthritis and an inadequate response to

methotrexate (Study Evaluating Rituximab's Efficacy in MTX iNadequate rEsponders (SERENE)). *Annals of the rheumatic diseases*. 2010;69. doi: 10.1136/ard.2009.119933

62 Genovese MC, Covarrubias A, Leon G, *et al.* Subcutaneous abatacept versus intravenous abatacept: a phase IIIb noninferiority study in patients with an inadequate response to methotrexate. *Arthritis and rheumatism*. 2011;63. doi: 10.1002/art.30463

63 Rubbert-Roth A, Tak PP, Zerbini C, *et al.* Efficacy and safety of various repeat treatment dosing regimens of rituximab in patients with active rheumatoid arthritis: results of a Phase III randomized study (MIRROR). *Rheumatology (Oxford, England)*. 2010;49. doi: 10.1093/rheumatology/keq116

64 Yang Y, Li J, Liu J, *et al.* Safety and efficacy of peficitinib in Asian patients with rheumatoid arthritis who had an inadequate response or intolerance to methotrexate: results of a multicenter, randomized, double-blind, placebo-controlled phase 3 study. *The Lancet Regional Health - Western Pacific*. 2024;42. doi: 10.1016/j.lanwpc.2023.100925

65 Taylor PC, Weinblatt ME, McInnes IB, *et al.* Anti-GM-CSF otilimab versus sarilumab or placebo in patients with rheumatoid arthritis and inadequate response to targeted therapies: a phase III randomised trial (contRAst 3). *Annals of the Rheumatic Diseases*. 2023;82. doi: 10.1136/ard-2023-224449

66 Tanaka Y, Kawanishi M, Nakanishi M, *et al.* Efficacy and safety of the anti-TNF multivalent NANOBODY® compound ozoralizumab in patients with rheumatoid arthritis and an inadequate response to methotrexate: A 52-week result of a Phase II/III study (OHZORA trial). *Modern Rheumatology*. 2023;33. doi: 10.1093/mr/roac119

67 Ye H, Liu S, Xu J, *et al.* Efficacy and Safety of CMAB008 Compared with Innovator Infliximab in Patients with Moderate-to-Severe Rheumatoid Arthritis Receiving Concomitant Methotrexate: A Randomized, Double-blind, Multi-center, Phase III Non-inferiority Study. *Rheumatology and Therapy*. 2023;10. doi: 10.1007/s40744-023-00544-2

68 Curtis JR, Stolshek B, Emery P, *et al.* Effects of Disease-Worsening Following Withdrawal of Etanercept or Methotrexate on Patient-Reported Outcomes in Patients With Rheumatoid Arthritis: Results From the SEAM-RA Trial. *Journal of Clinical Rheumatology*. 2023;29. doi: 10.1097/RHU.0000000000001893

69 Takeuchi T, Kawanishi M, Nakanishi M, *et al.* Phase II/III Results of a Trial of Anti–Tumor Necrosis Factor Multivalent NANOBODY Compound Ozoralizumab in Patients With Rheumatoid Arthritis. *Arthritis and Rheumatology*. 2022;74. doi: 10.1002/art.42273

70 Furst DE, Jaworski J, Wojciechowski R, *et al.* Efficacy and safety of switching from reference adalimumab to CT-P17 (100 mg/ml): 52-week randomized, double-blind study in rheumatoid arthritis. *Rheumatology (United Kingdom)*. 2022;61. doi: 10.1093/rheumatology/keab460

71 Liu Y, Liu S, Liu L, *et al.* Fine Comparison of the Efficacy and Safety Between GB242 and Infliximab in Patients with Rheumatoid Arthritis: A Phase III Study. *Rheumatology and Therapy*. 2022;9. doi: 10.1007/s40744-021-00396-8

72 Nasonov E, Fatenejad S, Feist E, *et al.* Olokizumab, a monoclonal antibody against interleukin 6, in combination with methotrexate in patients with rheumatoid arthritis inadequately controlled by methotrexate: efficacy and safety results of a randomised controlled phase III study. *Annals of the Rheumatic Diseases*. 2022;81. doi: 10.1136/annrheumdis-2021-219876

73 Kay J, Jaworski J, Wojciechowski R, *et al.* Efficacy and safety of biosimilar CT-P17 versus reference adalimumab in subjects with rheumatoid arthritis: 24-week results from a randomized study. *Arthritis Research and Therapy*. 2021;23. doi: 10.1186/s13075-020-02394-7

74 Behrens F, Koehm M, Rossmanith T, *et al.* Rituximab plus leflunomide in rheumatoid arthritis: A randomized, placebo-controlled, investigator-initiated clinical trial (AMARA study). *Rheumatology (United Kingdom)*. 2021;60. doi: 10.1093/rheumatology/keab153

75 Westhovens R, Rigby WFC, Van Der Heijde D, *et al.* Filgotinib in combination with methotrexate or as monotherapy versus methotrexate monotherapy in patients with active rheumatoid arthritis and limited or no prior exposure to methotrexate: The phase 3, randomised controlled FINCH 3 trial. *Annals of the Rheumatic Diseases*. 2021;80. doi: 10.1136/annrheumdis-2020-219213

76 Wiland P, Jeka S, Dokoupilová E, *et al.* Switching to Biosimilar SDZ-ADL in Patients with Moderate-to-Severe Active Rheumatoid Arthritis: 48-Week Efficacy, Safety and Immunogenicity Results From the Phase III, Randomized, Double-Blind ADMYRA Study. *BioDrugs*. 2020;34. doi: 10.1007/s40259-020-00447-6

77 Yamanaka H, Kamatani N, Tanaka Y, *et al.* A Comparative Study to Assess the Efficacy, Safety, and Immunogenicity of YLB113 and the Etanercept Reference Product for the Treatment of Patients with Rheumatoid Arthritis. *Rheumatology and Therapy*. 2020;7. doi: 10.1007/s40744-019-00186-3

78 Alten R, Batko B, Hala T, *et al.* Randomised, double-blind, phase III study comparing the infliximab biosimilar, PF-06438179/GP1111, with reference infliximab: Efficacy, safety and immunogenicity from week 30 to week 54. *Open Heart*. 2019;6. doi: 10.1136/rmdopen-2018-000876

79 Matucci-Cerinic M, Allanore Y, Kavanaugh A, *et al.* Efficacy, safety and immunogenicity of GP2015, an etanercept biosimilar, compared with the reference etanercept in patients with moderate-To-severe rheumatoid arthritis: 24-week results from the comparative phase III, randomised, double-blind EQUIRA study. *RMD Open*. 2018;4. doi: 10.1136/rmdopen-2018-000757

80 Smolen JS, Kremer JM, Gaich CL, *et al.* Patient-reported outcomes from a randomised phase III study of baricitinib in patients with rheumatoid arthritis and an inadequate response to biological agents (RA-BEACON). *Annals of the Rheumatic Diseases*. 2017;76. doi: 10.1136/annrheumdis-2016-209821

81 Chen X-X, Li Z-G, Wu H-X, *et al.* A randomized, controlled trial of efficacy and safety of Anbainuo, a bio-similar etanercept, for moderate to severe rheumatoid arthritis inadequately responding to methotrexate. *Clinical Rheumatology*. 2016;35. doi: 10.1007/s10067-016-3302-1

82 Takeuchi T, Yamanaka H, Tanaka Y, *et al.* Evaluation of the pharmacokinetic equivalence and 54-week efficacy and safety of CT-P13 and innovator infliximab in Japanese patients with rheumatoid arthritis. *Modern Rheumatology*. 2015;25. doi: 10.3109/14397595.2015.1022297

83 Smolen JS, Weinblatt ME, Van Der Heijde D, *et al.* Efficacy and safety of tabalumab, an anti-B-cellactivating factor monoclonal antibody, in patients with rheumatoid arthritis who had an inadequate response to methotrexate therapy: Results from a phase III multicentre, randomised, double-blind study. *Annals of the Rheumatic Diseases*. 2015;74. doi: 10.1136/annrheumdis-2014-207090

84 Genovese MC, van der Heijde DM, Keystone EC, *et al.* A phase III, multicenter, randomized, double-blind, placebo-controlled, parallel-group study of 2 dosing regimens of fostamatinib in patients with rheumatoid arthritis with an inadequate response to a tumor necrosis factor-alpha antagonist. *The Journal of rheumatology*. 2014;41. doi: 10.3899/jrheum.140238

85 Takeuchi T, Yamanaka H, Ishiguro N, *et al.* Adalimumab, a human anti-TNF monoclonal antibody, outcome study for the prevention of joint damage in Japanese patients with early rheumatoid arthritis: The HOPEFUL 1 study. *Annals of the Rheumatic Diseases*. 2014;73. doi: 10.1136/annrheumdis-2012-202433

86 Genovese MC, Han C, Keystone EC, *et al.* Effect of golimumab on patient-reported outcomes in rheumatoid arthritis: Results from the GO-FORWARD study. *Journal of Rheumatology*. 2012;39. doi: 10.3899/jrheum.111195

87 A Phase 2b/3, Randomized, Double-Blind Study Comparing Upadacitinib (ABT-494) to Placebo in Japanese Subjects With Moderately to Severely Active Rheumatoid Arthritis Who Are on a Stable Dose of Conventional Synthetic Disease-Modifying Anti-Rheumatic Drugs (csDMARDs) and Have an Inadequate Response to csDMARDs. Published Online First: 22 March 2016.

88 A Randomized, Controlled, Double-Blind, Parallel-Group, Phase 3 Study to Compare the Pharmacokinetics, Efficacy and Safety Between CT-P10, Rituxan and MabThera in Patients With Rheumatoid Arthritis. Published Online First: 23 May 2014.

89 A Randomized, Double-blind, Parallel, Placebo-controlled Study Assessing the Efficacy and Safety of Sarilumab Added to Non-biologic DMARD Therapy in Patients With Rheumatoid Arthritis Who Are Inadequate Responders to or Intolerant of TNF-α Antagonists. 2012.

90 Genovese MC, Fleischmann R, Kivitz A, *et al.* Efficacy and safety of sarilumab in combination with csDMARDs or as monotherapy in subpopulations of patients with moderately to severely active rheumatoid arthritis in three phase III randomized, controlled studies. *Arthritis Research and Therapy*. 2020;22. doi: 10.1186/s13075-020-02194-z

91 Coombs JH, Bloom BJ, Breedveld FC, *et al.* Improved pain, physical functioning and health status in patients with rheumatoid arthritis treated with CP-690,550, an orally active Janus kinase (JAK) inhibitor: results from a randomised, double-blind, placebo-controlled trial. *Annals of the rheumatic diseases*. 2010;69. doi: 10.1136/ard.2009.108159
